# Brief report: social media as a tool for scientific updating at the time of COVID pandemic

**DOI:** 10.1101/2020.06.12.20127555

**Authors:** Rita Murri, Francesco Vladimiro Segala, Pierluigi Del Vecchio, Antonella Cingolani, Eleonora Taddei, Giulia Micheli, Massimo Fantoni, COVID II Columbus Group

## Abstract

In the face of the rapid evolution of the COVID-19 pandemic, healthcare professionals on the frontline are in urgent need of frequent updates in the accomplishment of their practice. Hence, clinicians started to search for prompt, valid information on sources parallel to academic journals publications. Aim of this work is to investigate the extent of this phenomenon.

We administered an anonymous online cross-sectional survey to 645 Italian clinicians. 369 questionnaires were returned. 19,5% (n=72) of respondents were younger than 30 years-old; 49,3% (n=182) worked in Infectious Diseases, Internal Medicine or Respiratory Medicine departments, 11.5% (n=42) in Intensive Care Unit and 7.4% (n=27) were general practitioner. 70% (n=261) of respondents reported that their use of social media to seek medical information increased during the pandemic. 39.3% (n = 145) consistently consulted Facebook groups and 53.1% (n = 196) Whatsapp chats. 47% (n = 174) of respondents reported that information shared on social media had a consistent impact on their daily practice. In the present study, we found no difference in social media usage between age groups or medical specialties.

Given the urgent need for scientific update in face of the present health emergency, these findings may help understanding how clinicians access new evidences and implement them in their daily practice.

## Introduction

On March 30th, we attended the 8:30 am daily meeting of our recently born COVID Hospital. A talented colleague of mine took the microphone and read from a Whatsapp chat used only by medical professionals working in Northern Italy that they were observing a worrying number of pulmonary embolism cases. This was a new and previously unknown observation for us all. After the meeting, we came back to my ward where Mr A, a previously healthy man in his fifties recovering after a non-severe pneumonia due to SARS-CoV-2 was about to be discharged. He complained of a mild, yet worsening dyspnea and a lumbar pain. He had normal blood pressure and temperature values, cardiac rate was 100 bpm, respiratory rate 18 bpm, blood gases analysis showed pO2 87 mmHg, pCO2 34 mmHg and SatO2 96%. Keeping the recent meeting in mind, we decided to request a chest CT scan, that revealed massive pulmonary thromboembolism. Low-molecular-weight heparin (LMWH) at therapeutic dose was started, the patient improved within a few days and was then safely discharged. To what extent the recent informal communication lead to the diagnosis is difficult to define.

The first confirmed case of SARS-CoV-2 infection in Italy was identified in Rome on January 31th, 2020. Since then, the coronavirus disease 2019 (COVID-19) pandemic has spread around the world, catching many countries unprepared to face its enormous burden [1]. Moreover, even though SARS-CoV-2 primarily causes an interstitial pneumonia, it has been immediately clear that the clinical features of the disease could be unexpected and variable [2-3]. Hence, clinicians on the frontline started to search for rapidly available, valid information and to share, in turn, any relevant finding coming from daily practice or from preliminary data analysis.

## Methods

In order to investigate to what extent clinicians are seeking and using information coming from sources that are parallel to the scientific literature, we built an anonymous and voluntary questionnaire on SurveyMonkey. A total of 645 Italian clinicians potentially involved in the management of COVID-19 cases received the survey. Data were collected from the 5 ^th^ to the 14 ^th^ of April 2020 and analyzed from the 15 ^th^ to the 19 ^th^ of the same month. The present study was approved by the local Ethics Committee.

## Results

Three hundred sixty-nine questionnaires were returned. 19,5 % of respondents (n=72) were younger than 30 years-old and 10% (n=37) were more than 60 years-old; 21.9% (n=81) of the respondents worked in an Infectious Diseases department before the pandemic, 27.4% (n=101) in Internal Medicine or Respiratory Medicine, 11.5% (n=42) in Intensive Care Unit and 7.4% (n=27) were general practitioner. 57.5% of people (n=212) responded from Central Italy (including Lazio, our region), 30,4% (n=112) from Northern Italy and 10.6% (n=39) from Southern Italy. Fifty-two percent of respondents (n=191) were visiting patients with COVID-19 at least once per week, and 46.6% (n =172) visited confirmed COVID-19 cases every day. Data about how our colleagues sought information to obtain guidance for COVID-19 medical practice are presented in Table 1. Almost 80% of respondents (n=285) reported seeking information in peer-reviewed papers, yet an equal rate (78.4%; n=288) recurred to personal communications from colleagues working in other Centers at least twice per week, 39.3% (n=145) consistently consulted Facebook groups and more than the half (53.1%; n=196) reported to use Whatsapp chats for the same purpose at least once per week. Respondents characteristics are summarized in Table 1.

**Table 1.** Respondents characteristics

| Variable | Total sample<br>(N=369) | Social media impact<br>on clinical practice<sup>a,c</sup> | OR (CI) of reporting an<br>impact on clinical<br>practice<sup>d</sup> | P<br>Value |
| --- | --- | --- | --- | --- |
| Age |  |  |  |  |
| 20-29 | 72 (19.6) | 27 (48.2) | 0.05 (-0.51- 0.61) | .85 |
| 30-39 | 109 (29.6) | 52 (47.7) | 0.23 (-0.26-0.73) | .35 |
| 40-49 | 80 (21.7) | 45 (56.3) | 0.50 (-0.02-1.02) | .006 |
| 50-59 | 70 (19.0) | 37 (52.9) | 0.27 (-0.11-0.95) | .12 |
| 60+ | 37 (10.0) | 13 (35.1) | 0<sup>b</sup> |  |
| Position |  |  |  |  |
| Anesthesiologist/Intensive Care Unit | 42 (11.4) | 27 (64.3) | 0.70 (-0.40-1.82) | .21 |
| Surgeon | 49 (13.3) | 18 (36.7) | 0.25 (-0.821.34) | .64 |
| Pharmacist | 4 (1.08) | 2 (50.0) | 0.62 (-0.97-2.21) | .45 |
| Nurse | 3 (0.81) | 1 (33.3) | -0.04 (-1.83-1.74) | .96 |
| Infectious diseases specialist | 81 (22.0) | 38 (46.9) | 0.23 (-0.85-1.31) | .67 |
| Internal Medicine | 92 (25) | 36 (39.1) | 0.03 (-1.03-1.10) | .94 |
| Public Health doctor | 20 (5.43) | 10 (50.0) | 0.53 (-0.60-1.72) | .37 |
| Family doctor | 30 (8.15) | 24 (80.0) | 1.37 (0.23-2.59) | .02 |
| Pediatrician | 20 (5.43) | 6 (30.0) | 0.15 (-1.03-1.33) | .8 |
| Pneumologist | 10 (2.71) | 5 (50.0) | 0.24 (-1.07-1.56) | .72 |
| Psychiatrist | 4 (1.08) | 2 (50.0) | 0.52 (-1.08-2.14) | .52 |
| Radiologist | 6 (1.63) | 3 (50.0) | 0.42 (-1.03-1.88) | .57 |
| No position | 5 (1.35) | 2 (50.0) | 0<sup>b</sup> |  |
| Geographical Area |  |  |  |  |
| Northern Italy | 112 (30.4) | 49 (43.8) | -0.01 (-0.54-0.52) | .44 |
| Central Italy | 212 (57.6) | 104 (49.1) | 0.33 (-0.15-0.27) | .96 |
| Southern Italy | 39 (10.6) | 17 (51.5) | 0.43 (-0.68-1.55) | .21 |
| Frequency of COVID-19 cases' management |  |  |  |  |
| Never | 108 (29.3) | 44 (40.7) | -0.24 (-0.93-0.46) | .51 |
| Occasionally | 69 (18.7) | 32 (46.4) | -0.06 (-0.76-0.64) | .87 |
| Once a week | 19 (5.16) | 8 (42.1) | 0<sup>b</sup> |  |
| Everyday | 172 (46.7) | 90 (52.3) | 0.20 (-0.43-0.84) | .54 |

a. Survey question: "How impactful are the information acquired through social media for your daily practice?" answers: "Impactful and "Very impactful"
b. Set to zero because this parameter is redundant.
c. Missing data were not shown.
d. adjusted for age, position, geographical area and frequency of COVID-19 cases management

Seventy percent (n=261) of respondents reported that their use of social media to find medical information increased during the current pandemic. In terms of COVID-19 medical practice, information coming from social media were considered “enough” or “much” or “very much” useful by 82.9% (n=306) of the sample. To the question “During the last week, do you think that information shared on social media had an impact on your clinical practice for patients with COVID?” 28.7% (n=106) answered “enough” and 47.1% (n=174) “much” or “very much”.

## Discussion

In 2016, during the Zika epidemic, a protocol for data sharing during public health emergencies was issued by the World Health Organization [4]. Currently, several academic journals are trying to meet the instances of the medical community by hosting open-access COVID sections while speeding up their peer-reviewing process. Special pages have also been created to accelerate data sharing on this disease, such as the NEJM Coronavirus page [6], the Lancet COVID-19 Resource Centre [8], the BMJ’s Coronavirus (COVID) Hub [7], and the Cell Press Coronavirus Resource Hub [9]. Even scientific societies, foundations and consortia opened dedicated pages on their website [11]. Authors who decide to share useful information through special sections of academic journals could be accountable.

Our survey shows that, at the time of COVID pandemic, many clinicians react to their urgent need for updates by seeking information through unconventional sources instead of academic journals publications. Data obtained from colleagues working on different centers, Facebook groups and informal Whatsapp chats seem to be highly valued and trusted.

These findings may reflect the need of a more agile, user-friendly way to seek for medical information and updates, while the current epidemic is boosting the usage of social media to access to the complex, rapidly evolving amount of evidence that is increasingly emerging from all around the world. Interestingly, 150 responders (40.7%) reported to actively share medical information via social media “often” or “everyday”. This, on one hand, is coherent with the purpose of social media themselves but, on the other hand, it entails a broader shift in how professionals conceive the access to medical information, technological advances and scientific knowledge. We believe that it is important to acknowledge this phenomenon, taking into account its advantages in terms of rapidity, as well as the risk of spreading “fake news” or research exceptionalism, with potentially dangerous consequences for patients [5].

We strongly suggest that, during a pandemic, academic journals implement dedicated sections for rapid communications in the form of Forum sections, Rapid responses or Comments, and reserve peer reviewing for key points as needed [10, 12], in accordance to the cited WHO protocol for data sharing during public health emergencies [4]. Facilitated focus groups on social media could be another way to encourage discussion, even though, to our knowledge, no protocols are currently available.

Our study has several limitations. First, our sample was not uniformly distributed to all medical figures involved in COVID-19 epidemic, being intensive care doctors and primary care physicians likely underrepresented. Moreover, the results have to be interpreted with caution due to the small sample size. Finally, our findings about the impact of social media on clinical practice are based upon the personal perspective of the respondents.

## Conclusions

In conclusion, rapidly sharing information could have an invaluable impact during a pandemic such as that caused by SARS-CoV-2. Methods to promote a safe, open and rapid dissemination of relevant findings could provide a substantial benefit.

## Data Availability

The survey administered for the data collection of this work is available at the link below. All other data will be shared upon request.

https://it.surveymonkey.com/r/2BGGJVP

## Aknoledgements

* COVID II Columbus

Fabiana Baldi, Andrea Bellieni, Vincenzo Brandi, Maria Rosaria Calvello, Arturo Ciccullo, Antonella Cingolani, Camilla Cipriani, Giuseppe Maria Corbo, Cinzia Falsiroli, Massimo Fantoni, Enrica Intini, Giovanni Landi, Paolo Maria Leone, Rosa Liperoti, Francesco Macagno, Ilaria Martis, Giuliano Montemurro, Davide Moschese, Rita Murri, Cristina Pais, Giuliana Pasciuto, Annalisa Potenza, Sara Salini, Matteo Siciliano, Jacopo Simonetti, Eleonora Taddei, Matteo Tosato, Anita Tummolo, Francesco Varone, Giulio Ventura

---

**Figure 1.**
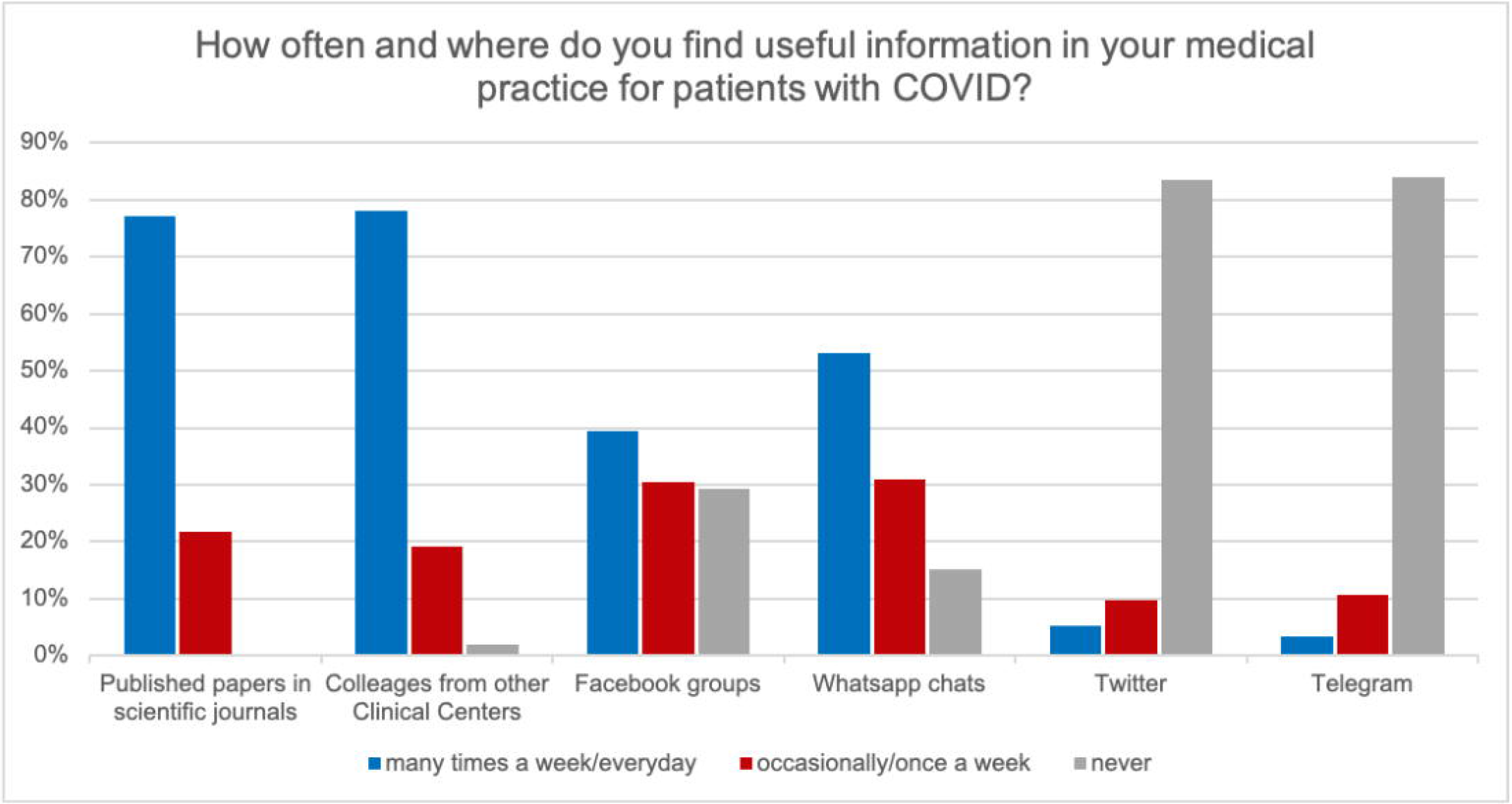
Questionnaire responses

